# Decoding the mechanophysiology for inhaled onset of smallpox with model-based implications for mpox spread

**DOI:** 10.1101/2025.06.17.25329814

**Authors:** Mohammad Yeasin, Mohammad Mehedi Hasan Akash, Abir Malakar, Azadeh A. T. Borojeni, Aditya Tummala, Jihong Wu, William D. Bennett, Wanda M. Bodnar, Julia S. Kimbell, Arijit Chakravarty, Julia R. Port, Saikat Basu

**Affiliations:** South Dakota State University, Department of Mechanical Engineering, Brookings, SD 57007, USA; Florida State University FAMU-FSU College of Engineering, Department of Mechanical Engineering, Tallahassee, FL 32310, USA; Harvard University, Paulson School of Engineering and Applied Sciences – Biomedical Engineering Program, Cambridge, MA 02138, USA; University of North Carolina School of Medicine, Center for Environmental Medicine, Asthma and Lung Biology, Chapel Hill, NC 27599, USA; University of North Carolina School of Public Health, Department of Environmental Sciences and Engineering, Chapel Hill, NC 27599, USA; Enthalpy Analytical, Durham, NC 27713, USA; University of North Carolina School of Medicine, Department of Otolaryngology/Head and Neck Surgery, Chapel Hill, NC 27599, USA; Fractal Therapeutics, Lexington, MA 02420, USA; Laboratory of Transmission Immunology, Helmholtz Centre for Infection Research, 38124 Braunschweig, Germany

**Keywords:** Inhaled virus transmission, Orthopoxviruses, Smallpox, Mpox, Respiratory fluid dynamics, Computational physiology, Critical exposure duration

## Abstract

Orthopoxviruses can transmit via inhalation of virus-laden airborne particulates, with the initial infection triggered along the respiratory pathway. Understanding the flow physics of inhaled aerosols and droplets within the respiratory tract is crucial for improving transmission mitigation strategies and elucidating disease pathology. Here, we introduce an experimentally-validated physiological fluid dynamics model simulating inhaled onset of smallpox caused by the variola virus of Orthopoxvirus genus. Using high-fidelity Large Eddy Simulations, we modeled airflow and particulate motion within anatomical airway domains reconstructed from medical imaging. By integrating these simulations with viral concentration and individual immune factors, we estimated critical exposure durations for infection onset to be between 1 − 19 hours, aligning with existing smallpox literature. To formalize the broader applicability of this framework, we extended our analysis to mpox virus, a circulating pathogen from same genus. For mpox, the mechanophysiological computations indicate a critical exposure window of 24 − 40 hours; however, this can vary significantly—from as short as 8 hours to as long as 127 hours—depending on virion concentration fluctuations within inhaled particulates, assuming happenstance of viral evolution. Predictably longer than the critical exposure durations for smallpox, the mpox findings still strongly suggest the possibility for airborne inhaled transmission during prolonged proximity.

## Introduction

Before eradication in the late 1970s through widespread vaccination efforts^1–3^, smallpox inflicted catastrophic mortality and morbidity on a global scale, claiming hundreds of millions of lives throughout history. The variola virus (VARV), which is the etiological agent responsible for smallpox, belongs to the Poxviridae family, with sub-family Chordopoxvirinae and the genus Orthopoxvirus, that also includes mpox virus, vaccinia virus, cowpox virus, and several other animal poxviruses which cross-react serologically^4^. The poxviruses consist of a single, linear, double-stranded DNA molecule ranging from 130 to 375 kilobase pairs and replicate within the cytoplasm of host cells. Under electron microscopy, they appear brick-shaped and typically measure approximately 300 by 250 by 200 nm (e.g., see Fig. 1A, with inset). Notably, the extensive body of research and resulting data on smallpox^5–9^ position it as *the* ideal model Orthopoxvirus for testing new mechanistic frameworks aimed at predicting its detailed transmission dynamics. These models may then be prospectively applied toward studying the disease onset mechanisms for other related (and, circulating) pathogens^10,11^. Among such examples, the mpox virus (MPXV)^12,13^ has recently prompted global health emergencies; notably declared as a Public Health Emergency of International Concern (PHEIC) by the World Health Organization (WHO)^14^ in 2022 and 2024, in view of its rapid international spread and potential to cause severe illness, particularly in immunocompromised individuals. Other poxviruses, such as cowpox and vaccinia, typically cause localized human infections; however, they do remain significant given their zoonotic potential and crucial role in vaccine development.

**Figure 1.**
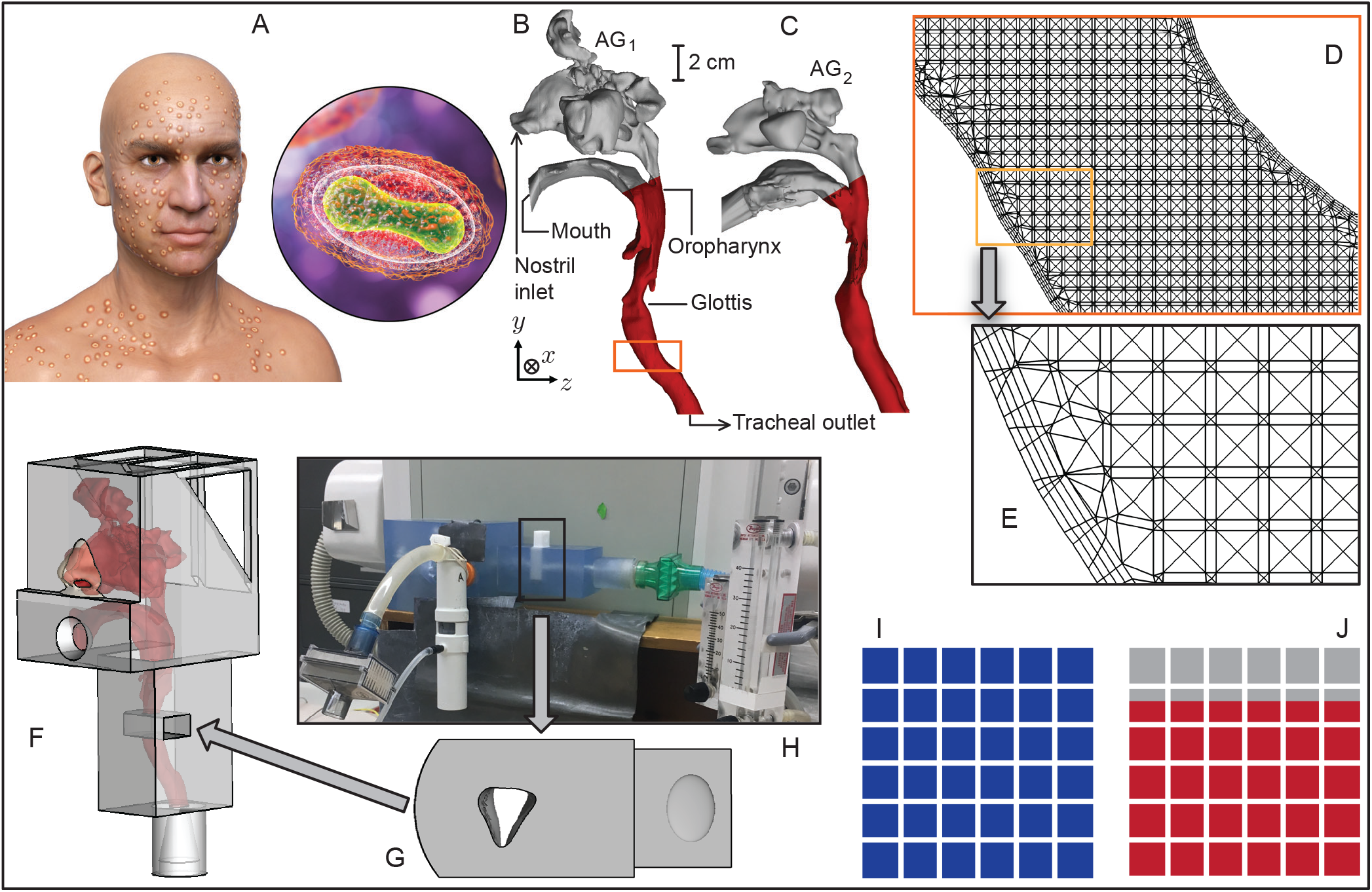
Mechanophysiological domain. Panel A depicts a cartoon illustration of a smallpox patient with pustules; the inset close-up shows the variola virus structure with outer membrane. Similar skin lesions can also be found in currently circulating viruses from the Poxviridae family, such as the mpox virus, which bears a similar virion morphology. The visual is adopted with a perpetual license agreement from the Getty Images^®^. Panels B and C respectively show the anatomical test airway domains (respectively labeled anatomical geometry 1 or AG_1_ and anatomical geometry 2 or AG_2_, built from high-resolution medical-grade computed tomography imaging). The red regions mark the initial infection trigger sites along the upper respiratory tract; downwind particulate penetration across the tracheal outlet to the lower airway is also tracked to account for bronchial infection expression. Panels D and E zoom on the orange box in B, demonstrating the mid cross-section for the mesh with 6.0 million tetrahedral cells, along with the four prismatic layers along airway walls. Panel F shows the 3D-printable digital model (derived from the AG_1_ reconstruction), with a scaled-up view of the separable glottic plug included in G. Panel H demonstrates the experimental setup for mimicking particulate transport using a nebulizer. The location where the glottic plug is inserted in the main 3D-printed structure is indicated within the black rectangle. Panels I and J compare the simulated (in blue) and experimental (in red) measurements for localized deposition fractions at the glottis. The colored area coverages are proportional to the inhaled glottic deposition fraction (with respect to the total number of particulates administered to the numerical space) for 9.5-*µ*m particulates. See Methods for details on the experimental benchmarking and validation. **Note:** the length scale for the anatomical domains is included between panels B, C.

Returning to VARV, its primary mode of transmission—beyond direct contact—involves the inhalation of virus-laden aerosols (typically ⪅ 5 *µ*m in diameter) and droplets (*>* 5 *µ*m) formed from an infected person’s respiratory secretions^5,15,16^. Consequently, a systematic understanding of how the inhaled airflow physics^17–20^ would influence the movement of the virus-bearing airborne particulates within the respiratory tract of an exposed subject is crucial toward elucidating the mechanism behind such infections. Be that as it may, although well-developed models exist for agent-based spatial transmission of smallpox in confined environments^6,21,22^, efforts on intra-airway exploration beyond understanding virus tropism and host immunity have been limited^23^. In particular, relatively little attention has been invested in elaborating the physical mechanics of virus propagation and the onset process of infection within the airway, as influenced by respiratory airflow streamlines. The VARV, when inhaled, primarily targets the mucosal epithelial cells at the oropharyngeal tissues—in the lower segment of the pharynx—along with spreading throughout the downwind tracheobronchial region of the respiratory system^6,24,25^. In such a perspective, to derive a physics-guided understanding of the infection onset process, this work aims to address the following mechanistic questions that arise from the complex interplay between the intra-airway dynamics of inhaled pathogen-bearing particulates and the viral biology:

Q_1_. What sizes of inhaled virus-laden particulates would preferentially deposit at the infective tissue sites along the airway, and how much of infectious viral load could they carry?

Q_2_. More poignantly, what could be the critical exposure duration that may lead to the onset of smallpox infection during airborne inhaled transmission?

To answer Q_1_ and Q_2_, this study develops high-fidelity computational fluid dynamics simulations of inhaled air and particulate transport within anatomically realistic respiratory domains (see Fig. 1B-E) built from medical-grade airway imaging data. As a validation exercise, the resulting findings are also benchmarked against representative physical experiments conducted within a 3D-printed anatomical cast (designed from one of the test geometries; see Fig. 1F-J). Ultimately, the flow physics-based trends assuming typical inhaled particulate size distribution are translationally integrated with viral concentration embedded in the inhaled particulates and individual immunological factors, for a precise quantification of the critical exposure duration that may trigger infection in an exposed individual. Therein, the established infectious dose of smallpox, typically between 10 − 100 plaque-forming units (pfu)^22,26–28^ and quantifying the minimum number of virions capable of triggering a new infection^29^, serves as a key cross-disciplinary parameter in determining the critical airborne exposure duration for infection onset.

The predictions for the critical exposure duration derived from our mechanics-guided inhalation analysis have been compared with the established exposure window estimates for smallpox transmission^6^. We do see a near-exact alignment between the exposure thresholds from intra-airway inhalation modeling and the known data, demonstrating the fidelity of our approach. Building on this, the study next explores the potential multifocal application of the in silico framework—using virus-specific data (such as the infectious dose and the viral concentration in host ejecta)—to evaluate the inhaled airborne transmission parameters for MPXV, a virus with similar morphology to the VARV. Specifically, MPXV shares 96.3% identity within the central region of the genome encoding essential genes, and 84.5% identity overall, with VARV^30^. Preliminary mechanophysiological findings from this work have been presented at the biofluid mechanics sessions of the 2024 Annual Meeting of the American Physical Society’s Division of Fluid Dynamics^31^ and at the 26^th^ International Congress of Theoretical and Applied Mechanics^32^.

## Results

### Intra-airway regional transmission trend as a function of inhaled particulate sizes

The test adult human respiratory tract geometries, hereafter referred to as anatomical geometry 1 or AG_1_ and anatomical geometry 2 or AG_2_, are depicted in Fig. 1B-C, showing their sagittal profiles. These geometries represent typical, disease-free airway cavity shapes in exposed individuals. They serve as the numerical domains for simulating the inhalation and transmission of virus-laden particulates, which can potentially lead to infection. Subsequently, Fig. 2A-B display the simulation-derived heatmaps for inhaled particulate deposition and penetration fraction, *η*_*k*_ (in %); it measures the net percentage of inhaled particulates that: (a) directly deposit at the infective tissue regions along the upper airway (colored red in Fig. 1B-C); and (b) penetrate through the tracheal outlet (see Fig. 1B) to move downwind into the infective bronchial airspace within the lower airway. These deposition and penetration fractions (with *η*_*k*_ summing up the two) are obtained as functions of the monodispersed particulate diameters *d ∈* [0.1, 50.0] *µ*m tested computationally. Considering the two modeled inhalation rates of 15 and 30 L/min (respectively mimicking relaxed and moderate breathing conditions^35^) in the two anatomical test geometries AG_1_ and AG_2_, the computational data comprises *k ∈* {1, 2, 3, 4}. In all geometry-flow combinations, *η*_*k*_ *≳* 30% for *d* ⪅ 9 *µ*m; the corresponding heatmap regions are highlighted within red boxes in Fig. 2A-B.

**Figure 2.**
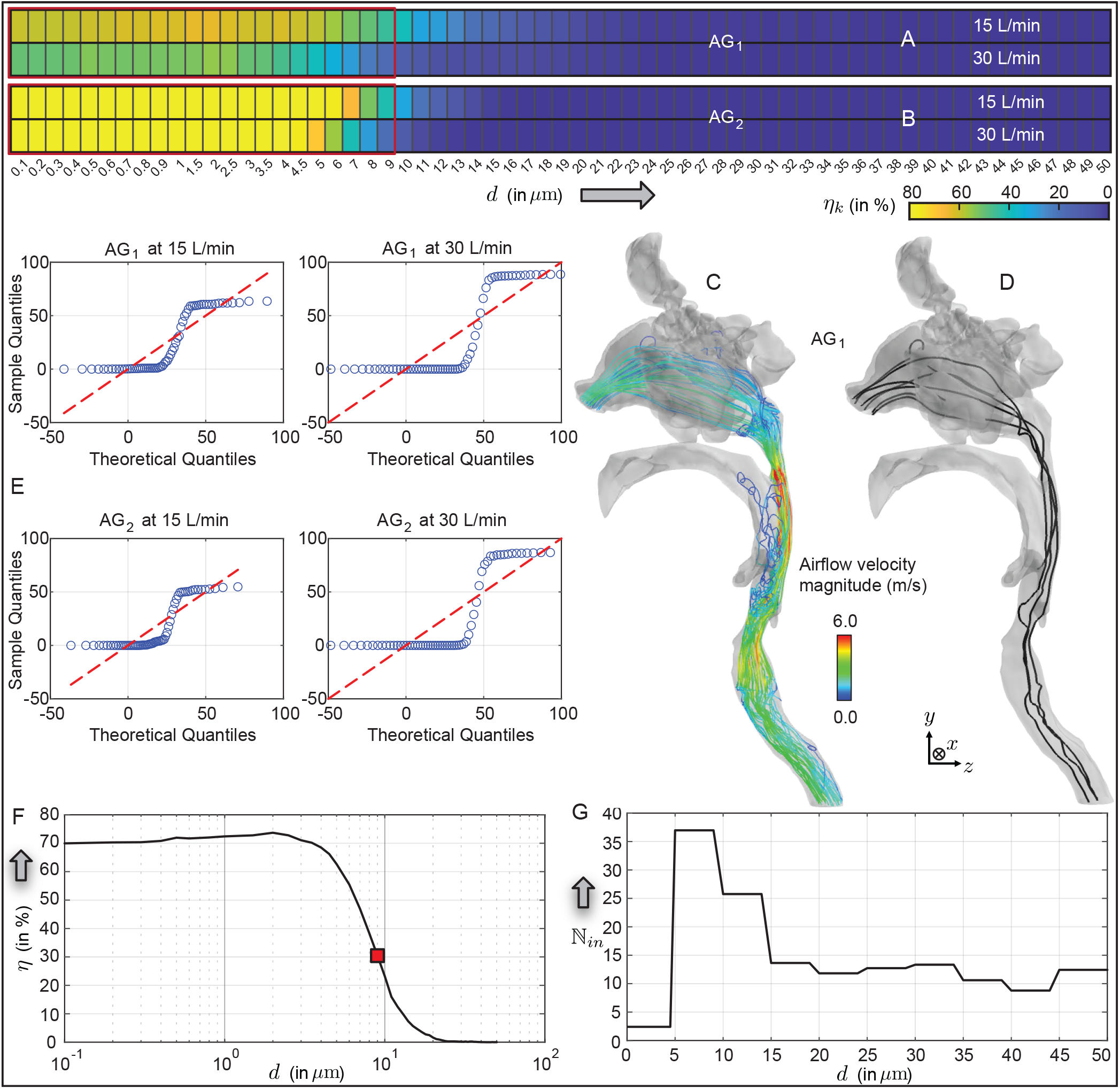
Simulated inhaled transport trend. Panels A and B respectively show the numerically simulated deposition and penetration percentage *η*_*k*_, summing the percentage of inhaled particulates that: (i) directly deposit at the infective tissue regions along the upper airway; (ii) penetrate downwind through the tracheal outlet into the infective bronchial airspace. The four rows in the heatmaps correspond to AG_1_ and AG_2_ (as marked), for inhalation rates 15 and 30 L/min. Considering all four geometry-flow combinations, *η*_*k*_ *≳* 30% for *d* ⪅ 9 *µ*m. The corresponding heatmap regions are highlighted within red boxes. Panel C shows 50 representative simulated airflow velocity magnitude streamlines (with 25 streamlines initiating from each nostril plane) during 15 L/min inhalation. Panel D depicts the simulated trajectories for 10 representative 1.5-*µ*m particulates; with 5 starting from each nostril plane. Panel E includes the Q-Q plots for the deposition and penetration trends as a function of the particulate sizes. Therein, the horizontal axis shows theoretical quantiles from a fitted normal distribution, the vertical axis shows sorted sample quantiles, and the red dashed line marks perfect normality. Panel F demonstrates the mean deposition and penetration rate *η* (in %), averaged across the test geometries and breathing rates. The red square marks the point when *η* first exceeds 30%. Based on the reported particulate size distribution in respiratory ejecta^33^, panel G shows the number of environmentally dehydrated^34^ particulates of each test size assumed to be inhaled per minute by an exposed subject.

Representative inhaled airflow streamlines and particulate trajectories are respectively shown in Fig. 2C-D. The simulations return the following inlet-to-outlet total pressure gradients (Δ*P*) based on the flow conditions (characterized by the inhaled airflux *Q*, in L/min): in AG_1_, Δ*P* = 31.91 Pa for *Q* = 15 L/min and Δ*P* = 106.11 Pa for *Q* = 30 L/min. In AG_2_, Δ*P* = 16.61 Pa for *Q* = 15 L/min and Δ*P* = 53.10 Pa for *Q* = 30 L/min. Herein, Δ*P* is measured as the absolute difference between the simulated pressure at the tracheal outlet and the mean of the inlet pressure values at the two nostrils in each geometry.

### Rank order testing to assess flow-geometric coupling and homogeneity in transmission trend

Within AG_1_ and AG_2_, the simulated deposition and penetration efficiencies (*η*_*k*_, in %) as a function of inhaled particulate diameters *d*, exhibit similar distribution shapes. This can be seen in the Q-Q (quantile-quantile) plot of each distribution against a normal distribution (note the separate panels in Fig. 2E), which also shows a strong deviation from normality. We formally tested the distributions for AG_1_ and AG_2_ for similarity, assessing the rank order correlation between them, using both Spearman’s and Kendall’s rank correlation coefficients, appropriate for non-normal distributions^36^. These tests involved ranking the inhaled particulate sizes (*d*) based on *η*_*k*_, and comparing the resulting orderings between AG_1_ and AG_2_. Table 1 presents the rank orders of *η*_*k*_ values as a function of the corresponding inhaled particulate sizes, comparing the simulated data from AG_1_ and AG_2_ at identical inhalation rates.

**Table 1.** Geometry-flow coupling. Rank order test data between AG_1_ and AG_2_ deposition fractions at the infective regions, taking combinations of the 15 L/min and 30 L/min simulation findings for *η*_*k*_ (in %), as function of the inhaled particulate diameters *d* (in *µ*m); see Fig. 2A-B.

| 15 L/min inhalation rate |  |  | 30 L/min inhalation rate |  |
| --- | --- | --- | --- | --- |
| | Correlation coefficient ($R$) | $p$-value | Correlation coefficient ($R$) | $p$-value |
| Spearman | 0.9193 | $2.1644 \times 10^{-26}$ | 0.9244 | $3.2144 \times 10^{-27}$ |
| Kendall | 0.8212 | $2.6557 \times 10^{-18}$ | 0.8605 | $1.3977 \times 10^{-18}$ |

The Spearman’s rank correlation coefficients were *R* = 0.9193 for 15 L/min inhalation and *R* = 0.9244 for 30 L/min inhalation, both with *p <<* 0.00001, indicating a highly significant monotonic relationship. Similarly, with the robust sample size of 63 data points (the count being the total number of particulate diameters tested for each inhalation rate; see the discrete values along horizontal in Fig. 2A-B) in each simulation, the Kendall rank correlation test yielded *R* = 0.8212 for 15 L/min and *R* = 0.8605 for 30 L/min, both with *p <<* 0.00001. These complementary results demonstrate a strongly consistent association between the rankings across the two geometries. Overall, the data reveals a highly reliable and statistically significant correlation in the ordering and monotonic trends of *η*_*k*_ values across different geometries and inhalation rates. The extremely small *p*-values (see Table 1) also reinforce the likelihood that the derived correlation measurements are genuine and not owing to random chances.

Backed by the consistent rank-ordering of *η*_*k*_ across the two test geometries for each inhalation rate and considering an equal mix of 15 L/min (for relaxed breathing) and 30 L/min (for moderate breathing) inhaled airflux in the exposed subject, we have subsequently obtained the averaged deposition and penetration fraction 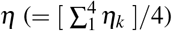 from the tested set of inhalation rates and geometries. This helps facilitate a generic streamlined trend of the inhaled transmission parameters for the remainder of this analysis. Serving as the primary computation-derived input to our study, *η* dictates inhaled transmission levels of virus-laden particulates to the infection-prone airway sites; the corresponding mean transmission rates are plotted in Fig. 2F.

### Temporal assessment of inhaled virus transmission

Figure 2G describes the size distribution of particulates N_*in*_(*d*), inhaled per minute—assuming they consist of dehydrated respiratory ejecta from an infected host. The data is adopted from established findings^33^ on the size distribution of aerosols and droplets emitted during normal speech and silent breathing. By coupling the inhaled size distribution N_*in*_(*d*) with the deposition and penetration efficiency *η*(*d*), it is straightforward to show that the inhaled liquid volume reaching the infective airway sites per minute would be:

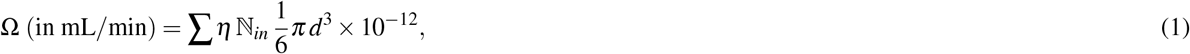

with the summation implying that equation 1 accounts for all particulate sizes that reach the infective airway sites and *d* representing the particulate diameters in *µ*m. Subsequently, considering the viral load carried by the liquid volume (as evaluated in equation 1), we can estimate the critical exposure duration for infection onset:

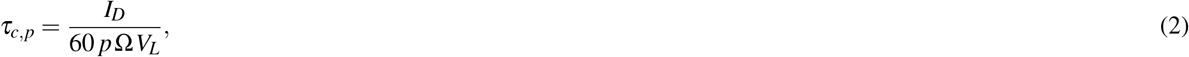

where *I*_*D*_ (in pfu) quantifies the infectious dose, *V*_*L*_ (in pfu/mL) is the mean viral loading within inhaled particulates, *τ*_*c,p*_ (in hours) is the critical exposure duration for infection onset, and *p*, as a measure of the dose-response relationship^38,39^, represents the potency (in %) of the plaque-forming units at triggering infection in an exposed subject. This is because while the plaque count (in pfu) quantifies the number of viral invasions within a monolayer of susceptible host cells, not all plaque-forming units are capable of initiating an infection in a real airway, which involves overcoming local and systemic mucosal defenses and immunological barriers. So, if the pfu potency is (say) *p*%, then out of every 100 infectious virions reaching the susceptible tissue sites, *p* of them could be presumed successful at invading the cells launching new infection.

### Critical exposure time for inhaled onset of VARV – with validation

Figure 3A depicts the trend for *τ*_*c,p*_ (or, in a simpler notation as in the plotted axis, *τ*_*c*_) for *I*_*D*_ *∈* [10, 100] pfu^22^. The upper trend lines correspond to lower values of *p* (implying fewer of the plaque-forming units ending up at the infective sites are successful at launching infection)—thereby lengthening *τ*_*c*_. We conservatively^40,41^ implement *p* (in %) *∈* [60, 100]. Figure 3B re-views the data from Fig. 3A, but from the perspective of *p* as the independent variable; here, the top-most and the bottom-most levels of the trend lines correspond, respectively, to the maximum and minimum *I*_*D*_. In addition, Table 2 lists a subset of numbers from the data plotted in Fig. 3A-B. Overall, as evidenced in Fig. 3 and also Table 2, *τ*_*c*_ varies over the range *≈* 1.12 − 18.59 hours, aligning well with current estimates for smallpox^6^. In fact, strikingly enough in context of prediction alignment, for *p* = 63% (red curve in Fig. 3A-B), we have *τ*_*c*_ *∈* [1.77, 17.70] hours (see fourth column in Table 2), while the critical exposure window from the well-established Wells-Riley model^42–44^ (considering exposure to a standard pathogen concentration in confined air) stands at 1.7 to 16.7 hours for a 63% probability of getting infected^6^.

**Table 2.**
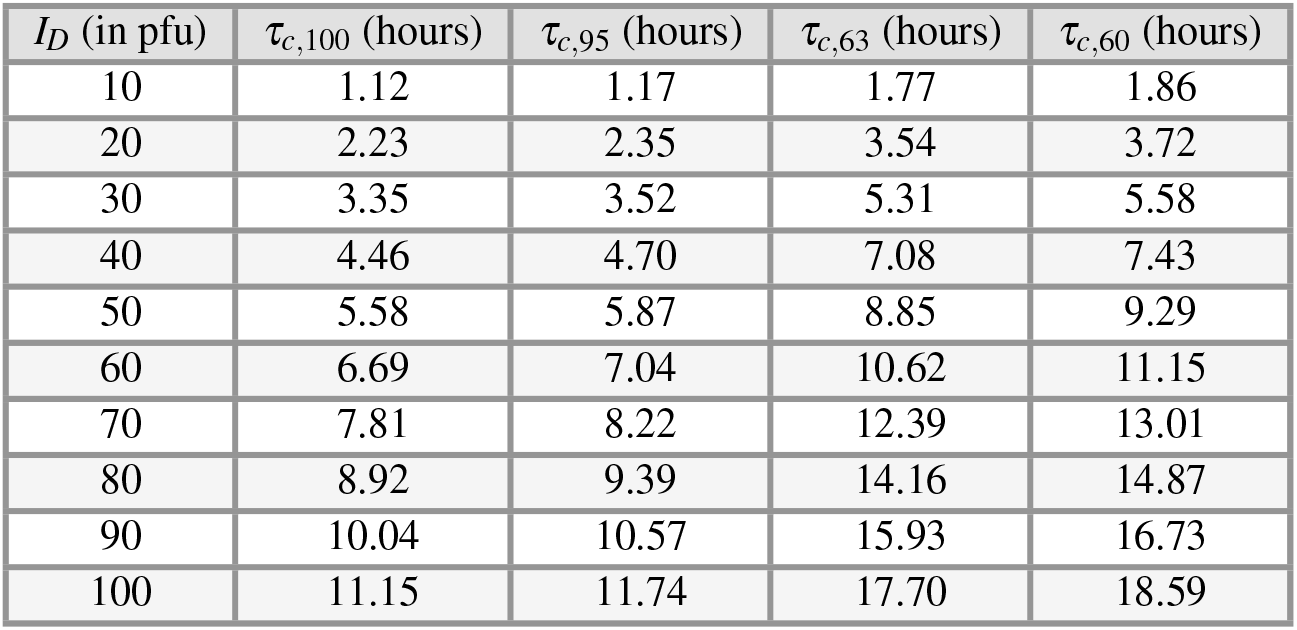
Explicit range for exposure window. Representative critical exposure durations (in hours) for *I*_*D*_ *∈* [10, 100] pfu. *τ*_*c,p*_ implies critical exposure duration assuming *p*% pfu potency at triggering infection inside an exposed human airway. Note that the listed numbers comprise a subset of the data plotted in Fig. 3A-B.

| $I_D$ (in pfu) | $\tau_{c,100}$ (hours) | $\tau_{c,95}$ (hours) | $\tau_{c,63}$ (hours) | $\tau_{c,60}$ (hours) |
| --- | --- | --- | --- | --- |
| 10 | 1.12 | 1.17 | 1.77 | 1.86 |
| 20 | 2.23 | 2.35 | 3.54 | 3.72 |
| 30 | 3.35 | 3.52 | 5.31 | 5.58 |
| 40 | 4.46 | 4.70 | 7.08 | 7.43 |
| 50 | 5.58 | 5.87 | 8.85 | 9.29 |
| 60 | 6.69 | 7.04 | 10.62 | 11.15 |
| 70 | 7.81 | 8.22 | 12.39 | 13.01 |
| 80 | 8.92 | 9.39 | 14.16 | 14.87 |
| 90 | 10.04 | 10.57 | 15.93 | 16.73 |
| 100 | 11.15 | 11.74 | 17.70 | 18.59 |

**Figure 3.**
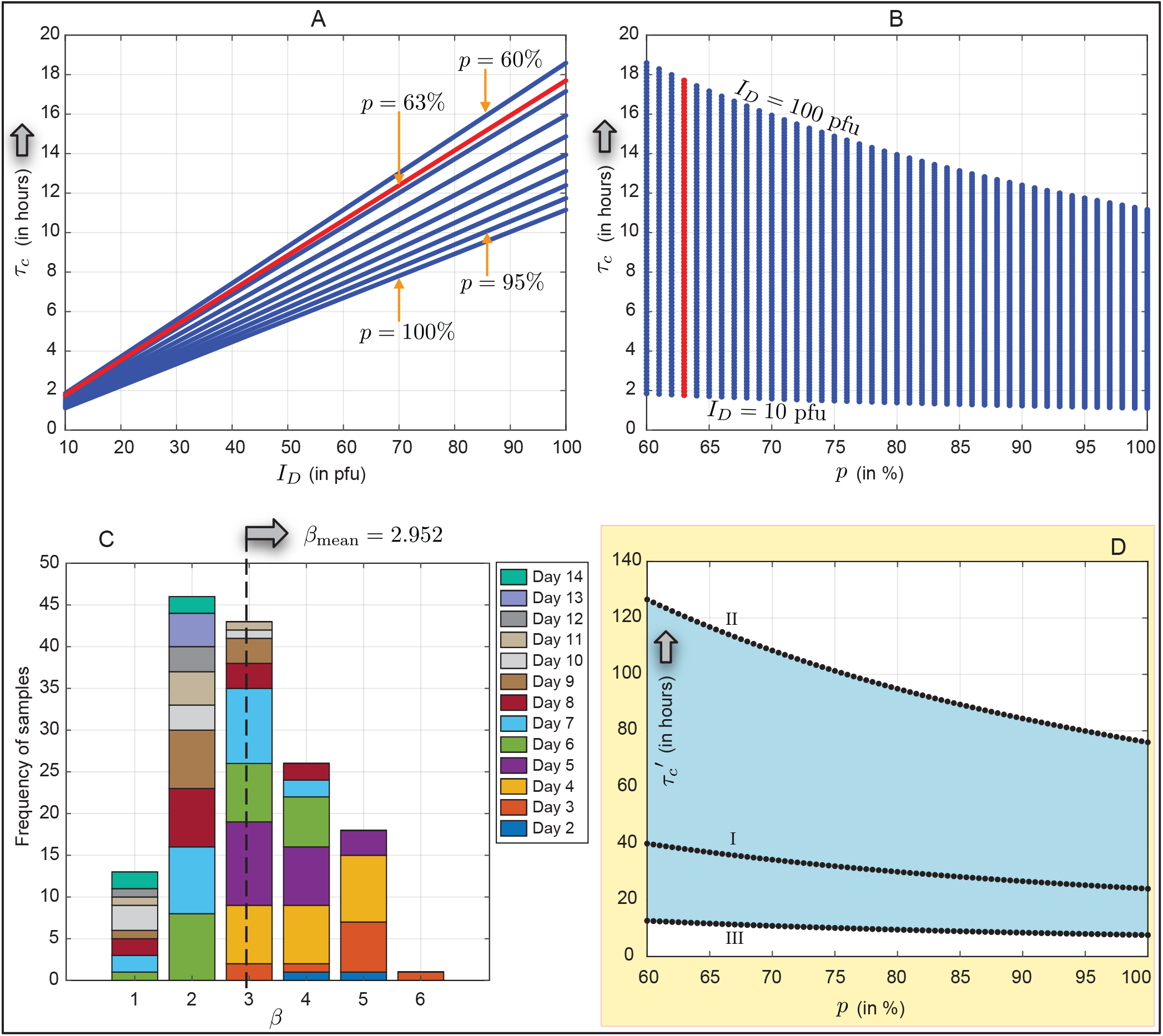
Exposure duration for infection onset (with Panels A − C for VARV; Panel D for MPXV) Panel A reports the critical exposure duration *τ*_*c*_ (in hours) for infectious dose *I*_*D*_ *∈* [10, 100] pfu and infectious virion potency *p ∈* [60, 100]%. The top trend lines are for lower *p*, implying longer time needed for infection onset (hence *τ*_*c*_ elevates). Panel B depicts the same data, but with *p* placed along the horizontal axis. Therein, each of the trend lines accounts for the entirety of the assumed *I*_*D*_ range, with the bottom-most point corresponding to *I*_*D*_ = 10 pfu and the top-most point corresponding to *I*_*D*_ = 100 pfu. In both A and B, the red curves are for *p* = 63%. Panel C records the sample frequency for *β*, where 10^*β*^ pfu/mL is the measured viral concentration in throat swabs, adopted from^8^. The data comprises 147 samples, collected from 32 patients, with a mortality rate of 34.38%. Explained on the right of panel C, the color code points to the day number during illness when the corresponding sample was collected. From this representation, the averaged index *β*_mean_ = 2.952 is used for calculating 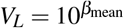 (in pfu/mL) while extracting the model projections. Panel D (in light yellow) highlights the mpox results on critical exposure duration 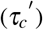. Curve I comprises findings with previously measured *V*_*L*_^37^; II shows the state if log_10_*V*_*L*_ reduces by 0.5; III shows the state if log_10_*V*_*L*_ increases by 0.5. The underlying blue region depicts the comprehensive 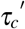 domain from viral concentration evolution, with perturbations on *V*_*L*_ following Δ log_10_*V*_*L*_ *∈* [−0.5, 0.5].

For the above calculation, we derived the viral loading *V*_*L*_ (in pfu/mL) by processing the virus titre data of throat swabs in 147 samples collected from 32 patients, reported in a seminal 1970s’ study^8^ (additionally, also see^9,45,46^). Figure 3C presents the sample frequency for *β*, where 10^*β*^ pfu/mL is the measured viral concentration. The averaged index *β*_mean_ = 2.952 is used in our analysis while deriving the *τ*_*c*_-trends in Fig. 3A-B.

### Model-based transmissibility of a related pathogen of same genus: Extension to mpox virus

With our model projections for critical exposure windows (*τ*_*c*_) for smallpox in agreement with existing data—we now extend the framework to analyze the airborne transmission characteristics of MPXV, investigating the ability of this pathogen to also spread through inhalation (i.e., without physical contact). As pointed out, MPXV is closely related to the VARV, with both belonging to the genus Orthopoxvirus. The minimum *I*_*D*_ for MPXV has been estimated to be 200 pfu^47^, with an average viral loading measured at *V*_*L*_ = 10^2.92^ pfu/mL^37^, based on Ct/CN counts from saliva samples. Inserting these values into equation 2 yields an estimated critical exposure duration 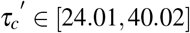 hours for inhaled onset of mpox; see curve I in Fig. 3D. The lower and upper bounds on 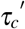 correspond to *p* = 100% and 60%, respectively. Such exposure windows could be plausible in a prolonged close-contact setting. The plausibility of MPXV transmission via the airborne inhalation route has implications for infection control (e.g., through effective ventilation and air change in confined spaces), as current guidelines are focused on fomites and larger respiratory droplets.

In addition, there are potential ramifications if viral evolutions were to occur in MPXV. As can be seen from equation 2, if the virus evolves such that the viral load in inhaled particulates (*V*_*L*_) changes, this may induce dramatic impacts on the airborne transmissibility of mpox, as it can significantly alter the length of time required for inhaling viral loads equivalent to *I*_*D*_, resulting in infection launch. For example, if we perturb the index on the viral loading value by Δ*β* = *±*0.5 (i.e., *V*_*L*_ = 10^2.92 *±* 0.5^), the 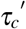 window fluctuates between 𝒪 (0) to 𝒪 (2) hours, with the precise model-based range being 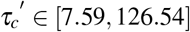 hours; bounded by the extrema on the curves III and II, respectively, in Fig. 3D. Especially at the lower end of the spectrum with reduced critical exposure requirements, the conditions can significantly enhance the airborne transmissibility of MPXV beyond prevalent norms, via inhalation of respiratory secretions expelled by infected hosts.

## Discussion

### On the significance of historical context while structuring the inhalation model

Our methodology mechanistically benefits from building upon the extensive body of smallpox research (the disease was formally declared eradicated by the WHO back in 1980^3^), enabling contextualization and cross-validation of historical data with modern computational methods. The general approach (with appropriate pathogen-specific inputs such as viral loading in host ejecta and the sites of infection origin in the exposed subject’s airway) can also contribute to the rapid assessment of transmission parameters for other Orthopoxviruses in the face of emerging outbreaks. The demonstrated extensibility underscores our framework’s potential as a versatile, physics-guided tool for swift risk evaluation based on detailed anatomical and inhalation fluid dynamics modeling.

Somewhat fortuitously, another “advantage” our model has benefited from is the lack of usable prior data on virion concentration in freshly expelled respiratory secretions from smallpox patients and, more generally, within their bodily fluids—specifically in units of DNA copies/mL. This scarcity is partly due to smallpox being no longer a circulating virus, and because genomic sequencing emerged only in the late 1970s^48,49^, coinciding with the eradication timeline of smallpox. Genomic techniques can be used to estimate viral concentration by analyzing the amount of viral nucleic acids present in a sample. However, for smallpox, such measurements are typically only available for purified historical viral preparations, as seen in^50^.

Nevertheless, the inhalation-based model requires viral concentrations in fresh respiratory secretions to estimate the inhaled viral load for an exposed individual. To address this, we utilized viral loading data expressed in pfu/mL (obtained from older studies involving swabs from contemporary patients^8^) within equations 1 and 2. This choice was also important for maintaining dimensional consistency within the mathematical framework, since the units for smallpox infectious dose data are also in pfu. In fact, the conversion between pfu and DNA copies for VARV remains unclear to this day^22^.

### On the implications from the test cohort size and the future directions

While the findings presented here are based on airflow-particulate transport patterns within only two anatomical respiratory domains, it is important to emphasize that this study does *not* aim to establish the statistical frequency of airborne transmissibility of the poxviruses. Instead, it simply seeks to evaluate the theoretical potential for such transmission within representative and realistic respiratory tracts, serving as a basis for understanding possible pathways rather than quantifying likelihood. Exploring how the variability in transmission dynamics might change as the geometric parameters of the airway regions are altered—owing to factors like age, health status, ethnicity or merely anatomical differences across individuals—would be a valuable direction for future research. Such investigations could help in assessing population-level risks and in tailoring more specific intervention strategies.

Regarding potential limitations, the geometric systems reconstructed from computed tomography (CT) scans are static and rigid, neglecting the dynamic compliance of airway tissues and tissue deformation that may occur during actual breathing in living subjects. These biomechanical factors can subtly influence airflow patterns, inhaled particulate trajectories, and deposition sites; these complex fluid-structure interactions that could be present in vivo are not incorporated into our modeling framework. Additionally, the simulated inhalation regimes did not account for mouth breathing, based on the rationale that oral inhalation accounts for less than 10% of breathing time in healthy adults^51^. While this simplification is reasonable for a holistic take on the typical breathing patterns, it may underestimate inhaled particulate deposition at the infective airway regions in scenarios where mouth breathing is more prevalent, such as during exercise or respiratory distress.

Several key questions also remain open. For example, a precise estimate of virion potency (*p*) in triggering new infections is still lacking, as current values are based on limited experimental data. More refined, virus-specific potency assessments could enhance the accuracy of predictions regarding critical exposure durations (by providing more specific inputs to equation 2). Furthermore, incorporating immunological factors—such as local mucosal defenses and individual innate and adaptive immune responses—into in silico models like the one proposed here remains an important future direction. Such enhancements could significantly improve the predictive power of this mechanophysiological framework and aid in the development of more nuanced public health strategies for controlling airborne pathogen transmission. The described modeling paradigm also assumes that both of the considered viruses (VARV and MPXV) have similar initial infection sites and tropism—a parameter which would benefit from future refinement once more detailed clinical data on MPXV clades becomes available.

### The main takeaway: On the airborne inhaled transmissibility of smallpox and mpox

The variola virus enters the body primarily through the respiratory pathway, triggering the initial smallpox infection within the mucosal epithelial cells along the airspace. Through high-fidelity, physiologically realistic computational fluid dynamics simulations of inhaled transport within CT-derived human respiratory tracts, integrated with virological parameters such as viral loading in host ejecta and established infectious dose levels, this study has yielded the following key insights into the airborne transmission of VARV: (a) smaller inhaled, virus-laden particulates are more effective at reaching the infective tissue sites along the airway; for instance, the relevant deposition and penetration efficiencies exceed 30% (of all inhaled particulates of the same size) for all particulate diameters ⪅ 9 *µ*m (see Fig. 2A-D and F); and (b) the critical exposure duration for transmission of viral load equivalent to the smallpox infectious dose to the infective intra-airway regions, thus resulting in inhalation-induced infection onset, is approximately between 1.1 to 18.6 hours (see Fig. 3A-B and Table 2). These projected durations align with existing literature^6,44^ that cites a critical exposure window of 1.7 to 16.7 hours (for a 63% probability of infection) based on the probabilistic Wells-Riley model for spatial transmission. Backed by representative experimental validation (described under Methods), the inferences in (a) and (b) above, thus, directly address the questions Q_1_ and Q_2_ posed in the Introduction.

Given that exposure to host-ejected respiratory particulates constitutes the primary (non-contact) transmission mode for Orthopoxviruses, it is important to distinguish our framework from existing transmission models^22^. While our study conceptualizes physiologically representational inhalation dynamics effects for intra-airway deposition and penetration of virus-laden particulates leading to a precise quantification of disease transmission parameters, most current models generally rely on agent-based, probabilistic methods to estimate viral exposure within confined external spaces, sometimes with room-level computational modeling of spatial pathogen distribution. The potential extensibility of our micro-scale framework is further demonstrated by its application to MPXV (from the same Orthopoxvirus genus), with the suggestion that its critical exposure duration could be substantially longer (roughly 24 to 40 hours), compared to that of smallpox. Prolonged though it may be, the projected time window still implies that respiratory aerosol transmission of mpox could be plausible in specific close-contact settings. For instance, the mpox critical exposure window could decline to as short as 8 hours, if the virion concentration in the inhaled pathogenic particulates fluctuates assuming the happenstance of viral evolution where the virus adapts to higher virus loads in the host airway and the infectious agents manage to become airborne by embedding in respiratory ejecta. Such pointed insights can inform public health policies on mpox, particularly for risk assessments of airborne transmission.

## Methods

### Airway geometry reconstruction from medical imaging

The test respiratory tract geometries AG_1_ and AG_2_ (their sagittal profiles shown in Fig. 1B-C), with disease-free airway cavity shapes, were reconstructed from de-identified high-resolution medical-grade CT imaging of adult human airways. The process entails radio-density thresholding from −1024 to −300 Hounsfield units^52^, along with selective manual editing of specific pixels for anatomical precision, to accurately capture the airspace from the CT slices in the DICOM (Digital Imaging and Communications in Medicine) format. To prepare the domains for numerical simulations of intra-airway inhaled transport, a representative grid refinement analysis was conducted on AG_1_, with the cavity being spatially segregated into approximately 4.0, 5.0, 6.0, 7.0, and 8.0 million graded, unstructured, tetrahedral elements, along with four layers of pentahedral cells (with 0.025-mm height for each cell and an aspect ratio of 1.1) extruded at the airway cavity walls to resolve the near-wall particulate dynamics; e.g., see Fig 1D-E. While the CT images were segmented on the image processing software Mimics Research 18.0 (Materialise, Plymouth, Michigan), the subsequent spatial meshing of the digitized stereolithography domains was carried out on ICEM-CFD 2023 R1 (ANSYS Inc., Canonsburg, Pennsylvania). Across the five progressively refined meshes (for AG_1_), we further assessed the flow resistance *R* (in Pa.min/L) to the simulated inhaled airflow field (moving at 30 L/min, measured at the outlet), calculated as |Δ*P*| */Q*, where |Δ*P*| in Pa represents the inlet-to-outlet total pressure gradient driving the airflow and *Q* denotes the volumetric airflux in L/min. The flow outcome sensitivity findings, as a function of the grid refinements, show:

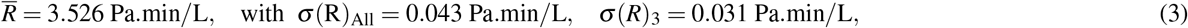

where the bar implies arithmetic mean, *σ* (·)_All_ denotes the standard deviation across all five grids, and *σ* (·)_3_ denotes the standard deviation of the simulated data from the last 3 grids (i.e., cases with 6.0, 7.0, and 8.0 million unstructured tetrahedral cells).

Based on the asymptotic convergence trend observed in the final three cases (see equation 3), the simplest 6-million-cell grid resolution was selected as the mesh density standard for the overall study. Consequently, the mesh used for the comprehensive analysis in AG_1_ comprised approximately 6 million tetrahedral elements. With the airspace volume in AG_1_ being 1.5 times larger than that in AG_2_, the appropriate number of cells for AG_2_ is estimated to be 6/1.5 = 4 million. To ensure adequate resolution, we conservatively adjusted the AG_2_ mesh to contain slightly more cells than this estimate, resulting in precisely 4.8 million tetrahedral elements. These spatial refinements are consistent with similar systems involving grid convergence and computational stability when simulating physiologically realistic airflow and particulate deposition within adult healthy human respiratory tracts (e.g., see^53,54^).

### Computational simulation of inhaled transport within the respiratory system

#### Mathematical framework for inhaled airflow simulation

Inhalation airflow patterns^35^ for relaxed (15 L/min) and moderate breathing (30 L/min) were replicated in the discretized test geometries using the Large Eddy Simulation (LES) scheme^55,56^, which explicitly resolves field eddies larger than the grid scale. Smaller fluctuations, known as subgrid scales, were filtered out, with their effects on larger scales approximated through the dynamic subgrid-scale kinetic energy transport model^57–59^. With incompressible and isothermal flow conditions for the inhaled air, the filtered continuity and Navier–Stokes equations were expressed as follows:

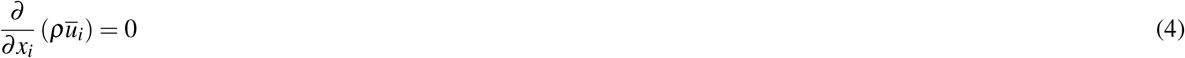

And

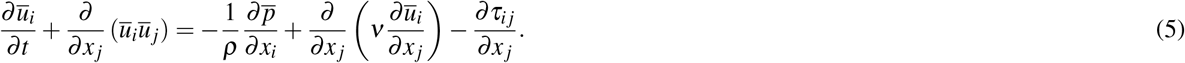

In these equations, 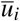 denoted the filtered (resolved) velocity component, while 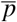 represented the filtered pressure. The parameters *ν* and *ρ* stood for the kinematic viscosity and the density of the inhaled, warmed air, respectively. The subgrid-scale (SGS) stress tensor, *τ*_*ij*_, was defined as

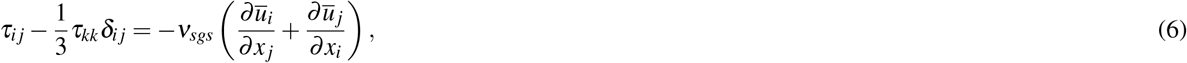

with *ν*_*sgs*_ representing the SGS kinematic viscosity and *δ*_*ij*_ being the Kronecker delta. The isotropic component of the SGS stresses, *τ*_*kk*_, was not explicitly modeled; instead, it contributed to the filtered static pressure. The instantaneous velocity field, incorporating SGS fluctuations, was then expressed as

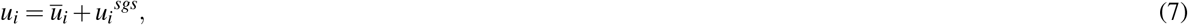

where 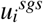 signified the SGS velocity fluctuations. The flow dynamics and particulate dispersion within the human respiratory system would be significantly influenced by the secondary flows, which are common in such complex geometries, as well as by the transition between laminar and turbulent regimes.

To accurately capture both transitional phenomena and secondary flow structures, we invoked the dynamic subgrid-scale kinetic energy transport model^57–59^. Within this framework, the SGS kinematic viscosity *ν*_*sgs*_ was computed based on the Kolmogorov-Prandtl hypothesis^60^ as:

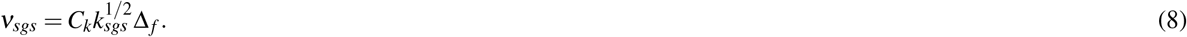

Here *C*_*k*_ was a model constant, and Δ _*f*_ was the filter size, defined as the cube root of the grid cell volume, Δ _*f*_ *≡* (cell volume)^1*/*3^. Also, the SGS kinetic energy, *k*_*sgs*_, could be mathematically represented as

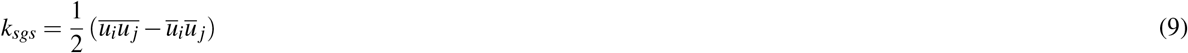

Subsequently, the spatial and temporal evolution of *k*_*sgs*_ was governed by the following filtered transport equation:

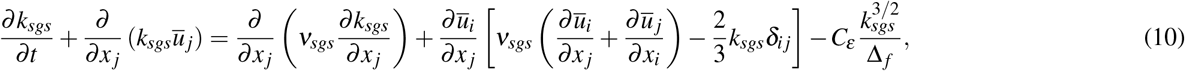

where the model constants *C*_*k*_ and *C*_*ε*_ were determined dynamically^57^.

The inhalation simulations applied no-slip boundary condition (i.e., zero velocity) at the airway walls, with the flow conditions being driven by pressure gradients. Therein, the numerical time steps were maintained at 0.0002 s for a total flow time of 0.35 s, consistent with reported findings^59^ regarding the time-step size necessary to fully resolve the unsteady turbulent airflow field in a realistic airway. Performed on a segregated solver with SIMPLEC pressure-velocity coupling and second-order upwind spatial discretization, the transient formulations used a bounded second-order implicit scheme, balancing between accuracy (owing to the second-order formulation), stability (from the implicit approach), and boundedness (to prevent non-physical oscillations). To monitor solution convergence, we minimized the mass continuity residual to 𝒪 (10^−4^) and the velocity component residuals to *O*(10^−5^). Additionally, assuming that inhaled air warms as it travels through the complex respiratory pathway, the air density *ρ* was set at 1.204 kg/m^3^ and its kinematic viscosity *ν* was 15.16*×*10^−6^ m^2^/s.

#### Mathematical framework for inhaled particulate tracking

The spatiotemporal evolution of inhaled particulates entering the airway through the nostril inlets was numerically tracked against the simulated airflow field. Considering dilute conditions for the resulting particulate dispersion, the momentum transfer was one-way coupled; i.e., the airflow continuum was assumed to impact the particulate motion, while the particulates had no effect on the underlying flow field. A Lagrangian-based inert discrete phase model, employing a Runge-Kutta solver, was utilized to numerically integrate the particle transport equation:

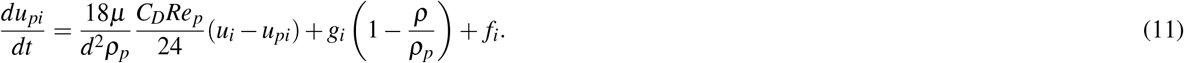

In this formulation, *u*_*pi*_ denoted the particulate velocity, *ρ*_*p*_ was the material density of the particulates, and *d* (as before) represented the tracked particulate diameter. The term *Re*_*p*_ referred to the particle Reynolds number, while *g*_*i*_ indicated the gravitational acceleration component in the *i*-direction. The coefficient *C*_*D*_ corresponded to the drag coefficient, and *f*_*i*_ represented additional body forces per unit particulate mass, such as the Saffman lift force exerted by the local flow shear on small particulates transverse to the airflow. Also, the inhaled particulates were assumed to be sufficiently large for Brownian motion effects to be negligible.

The drag contribution in equation 11 was evaluated using the following forms for *Re*_*p*_ and *C*_*D*_:

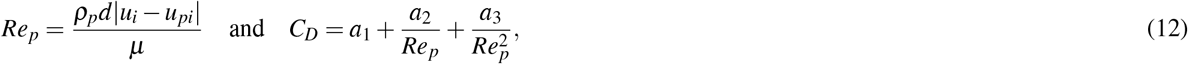

where *µ* denoted the molecular viscosity of the ambient air, and *a*_1_, *a*_2_, and *a*_3_ were functions of *Re*_*p*_, determined based on the spherical drag law^61^. Subsequently, the particulate trajectories were derived from their spatiotemporal locations, *x*_*i*_(*t*), obtained through the numerical integration of the following velocity vector equation:

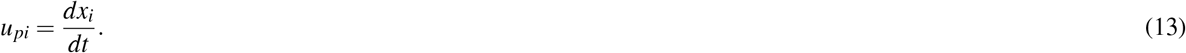

The tracked particulates were designed to mimic environmentally dehydrated respiratory ejecta from an infected individual, now being inhaled by an exposed subject. When expelled, liquid-based respiratory particulates generally lose water and decrease in size^34,62^, with the degree of shrinkage partly influenced by the proportion of non-volatile components within the particulates, such as dehydrated epithelial cell remnants, white blood cells, enzymes, DNA, sugars, and electrolytes. Therefore, while sputum is made up of up to 99.5% water, the post-dehydration particulates that are about to be inhaled have been shown to attain a density as high as 1.3 g/mL^34^, which is what has been enforced in the intra-airway particulate tracking simulations. The dehydration resulting in such density variation causes the diameter of an emitted particulate to shrink to 30% of its original size^34^, before being inhaled by the exposed subject. In other words, for example, an inhaled 1-*µ*m particulate would carry the same viral load embedded in its pre-dehydration 3.33-*µ*m size. The computationally tracked (inhaled) particulates, which mimicked the dehydrated and shrunken ejecta as described, were monodisperse and had the following diameters *d*: 0.1 − 0.9 *µ*m (with increments of 0.1 *µ*m); 1.0 − 4.5 *µ*m (with increments of 0.5 *µ*m); and 5.0 − 50.0 *µ*m (with increments of *1*.*0 µ*m)—beyond this size, prompt gravitational sedimentation is expected^34^, thus barring such particulates from being inhaled into the airway by the exposed subject. For each test diameter within the stated range, the total number of particulates tracked were *N*_*i*_: 1372 in AG_1_ and 2506 in AG_2_, with *N*_*i*_ being the number of surface elements in the mesh layer mapping the nostril inlets. The intra-airway spatial tracking of a particulate pathline was terminated once it entered the mesh element layer adjacent to the enclosing cavity walls. The particulates leaving the numerical domains through the tracheal outlets (see Fig. 1B-C) were also recorded; the finding represented the fraction of the inhaled particulates that were able to navigate downwind to the infective bronchial recesses in the lower respiratory tract^56^.

### Validation of the computational scheme by benchmarking with physical experiments

#### 3D printing of an anatomically realistic respiratory cast

To assess the numerically predicted particulate deposition trend vis-à-vis physical experiments, we generated a 3D-printed solid twin of AG_1_ (see Fig. 1F-H), made from the stereolithography material Watershed^®^ (DSM Somos^®^, Elgin, Illinois); the printing being done at ProtoLabs (Maple Plain, Minnesota)^63^. In the glottic region of the airspace (see marked region in Fig. 1B), we incorporated a hollow groove (see Fig. 1F) designed to snugly accommodate a plug (see Fig. 1G) that captures the CT-based internal anatomical topology at the glottis. The primary objective was to collect wall deposits within the plug, measure the localized deposition fraction, and compare these experimental measurements with our computational projections for localized deposition at the same region. The glottic plug was 3D-printed using an Original Prusa i3 Mk3 printer, produced by Prusa Research^®^, with flexible TPU filament supplied by SainSmart^®^.

#### Experimental logistics and measurements

Figure 1H shows the 3D-printed AG_1_ fitted to a negative pressure pump that drew a constant air flux of 30 L/min through the tracheal outlet for a period of 2 minutes before and during nebulization. To obtain intra-airway particulate transport trends, aerosolized droplets with a diameter of 9.5 *µ*m^64^ were delivered via a modified Pari LL jet nebulizer inserted at the mouth opening of the anatomical reconstruction. Each experimental run utilized 2 mL of a nebulized aqueous caffeine solution at a concentration of 10 mg/mL, with a total of three runs conducted. Following nebulization, we measured localized deposition after the droplets settled on the inner walls of the glottic plug (Fig. 1G) using ultraviolet (UV) absorbance of the caffeine solutions, analyzed with a Thermo Scientific Biomate 5 spectrophotometer at the University of North Carolina’s Biomarker Mass Spectrometry Core Facility. For each of the three samples, the glottic plug was rinsed in 60 mL of deionized water for 10 minutes using an ultrasonic bath. The calibration range was adjusted to bracket the samples, and we recorded the signal for each sample in triplicate. A blank test insert was rinsed in the same manner to ensure there was no background signal at 273 nm, using a 1 mL aliquot for the UV absorbance measurement. The resulting measurements are presented in Table 3, with *C*_*E*_ quantifying the caffeine concentration in the solutions resulting from rinsing the glottic deposits with deionized water, and *G*_*E*_ represents the percentage of the nebulized solution that landed in the glottis while inhalation was simulated by the pressure pump drawing in air.

**Table 3.** Validation test. Comparative data for glottic deposition from representative experiments and benchmarked computational simulations. See Fig. 1F-J for experimental setup and data visuals.

| Experimental measurements |  |  | Computational measurements, for validation |  |
| --- | --- | --- | --- | --- |
| Run # | $C_E$ (in $\mu\text{g/mL}$) | $G_E$ (in %) | $d_C$ (in $\mu\text{m}$) | $G_C$ (in %) |
| I | 4.014 | 1.205 | 9.25 | 1.638 |
| II | 4.301 | 1.291 | 9.50 | 1.797 |
| III | 4.001 | 1.201 | 9.75 | 1.374 |
| Mean | 4.105 | 1.232 | Mean | 1.603 |

Given the described experimental parameters, the primary concentration measurements *C*_*E*_ (in *µ*g/mL) from rinsing the glottic plug are utilized to derive the local deposition fractions *G*_*E*_ using the following conversion:

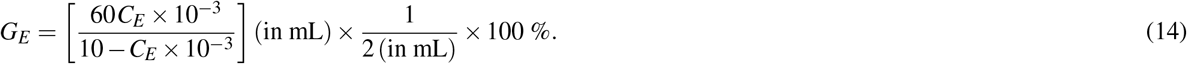

#### Benchmarked computational simulation for comparison with experimental measurements

To establish the reliability of the broader computational results, we conducted a benchmarking exercise by replicating the experimental conditions in silico, utilizing the same numerical flow scheme as implemented in the larger study. For a simulated airflow rate of 30 L/min in AG_1_, we computationally tracked particulates with diameters *d*_*C*_ *∈* 9.50 *±* 0.25 *µ*m (the perturbation on *d*_*C*_ accounting for fluctuations in the monodispersed nature of the nebulized particulates in the experiments). The material density of the simulated inert particulates was set at 1 g/mL, to mimic the aqueous solution used in the experiments. Similar to the nebulized droplets in the experiments, the simulated particulates were administered into the airspace, specifically for this benchmarking, through the mouth opening with an initial speed of 10 m/s (based on reported data on jet nebulizers)—the velocity vector lying on the *yz* plane and oriented at 45^*°*^ to both *y* and *z*-axes (see Fig. 1B)—ensuring a realistic entry angle through the sectional plane at the mouth. The last two columns in Table 3 report the computationally simulated deposition fractions at the glottis (the precise region marked by the experimental glottic plug); with the total number of particulates administered and tracked for each *d*_*C*_ being 1892, corresponding to the number of grid cells mapped across the planar cross-section at the mouth opening. These localized deposition fractions (*G*_*C*_) are equivalent to the ratio of the number of particulates depositing at the glottis over the total number of particulates administered, converted to percentage.

The mean glottic deposition fraction from the experiments (*G*_*E*_) is found to be approximately 0.8 times that of the computational projection (*G*_*C*_); see Table 3, and additionally Fig. 1I-J for a visual comparison. We argue that this comparability between the experiments and simulations could be deemed satisfactory, with the difference attributable to post-deposition surface displacement of the deposited droplets in the experimental plug owing to gravity, as well as to airflow-induced shear over the 2-minute duration of the experimental flux. There could also be loss of deposited liquids as the plug was removed from the main 3D print and transferred to deionized water bath for spectroscopic measurements. Note that dislodging deposited particulates from the inner walls of the 3D-printed plastic cast is easier than in a real airspace, where the enclosing tissue surfaces are typically infused with mucus and ciliary envelopes.

#### Virological inputs to the flow physics model

VARV, the causative agent of smallpox, is a classic example of an Orthopoxvirus, with an established infectious dose (*I*_*D*_) ranging between 10 − 100 pfu^22^; this range is used in equation 2. Critically though, for variola, the conversion of pfu to DNA copies is still an unresolved question. Earlier findings^8^ reported plaque counts from throat samples of smallpox patients (with variola major), enabling an estimation of the viral load distribution in particulates expelled by a host; accordingly, we have calculated the mean viral load *V*_*L*_ = 10^2.952^ pfu/mL (as an input to equation 2). The raw data comprised 147 throat swab specimen taken from 32 hospitalized patients at progressive stages of the disease over a 2-week period; see Fig. 3C. In the historical study^8^, the cotton swabs were soaked in Hanks’ basal salt solution (BSS) containing 0.5% bovine albumin and antibiotics (penicillin and streptomycin); subsequently, the virus titre was obtained from the fluid obtained by squeezing the swabs after they had been dipped in 1-mL of BSS. Although swabs can possibly carry higher pathogen concentration (than in exhaled particulates by a host) owing to the invasive nature of sample collection, the BSS dilution explains the standard use^22^ of the eventual titre information as the viral loading parameter in the airborne particulates (composed of dehydrated respiratory ejecta from an infective host) inhaled by an exposed subject.

For MPXV, the input value for *I*_*D*_ in equation 2 was set to 200 pfu, based on measurements for the Zaire V79-I-005 strain^47^. Additionally, the mean viral load (*V*_*L*_ = 10^2.92^ pfu/mL) was estimated from Ct/CN counts in saliva samples^37^ for MPXV belonging to clade II (subclade IIB)^65–67^. Herein, we note that the findings for MPXV could be further improved with matched data sets for each clade, which could additionally address clade differences in disease onset mechanisms. In general, for both Orthopoxviruses considered here, the reported range for the critical exposure duration (*τ*_*c*_) spans several hours: approximately 1 − 19 hours for VARV and 24 − 40 hours for MPXV; the latter based on current phylogenetic data. This assumes continuous exposure to a live pathogen source—and as such—while pox viruses like MPXV can indeed stay stable for hours^68^, the described mechanophysiological model remains agnostic to virus stability-related long-term inactivation of infectious particulates.

## Data Availability

All data produced in the present study are available upon reasonable request to the authors

## Acknowledgments

This work is primarily supported by S.B.’s National Science Foundation CAREER Award (Grant No. CBET 2339001). Supplemental support for 3D printing came from a pilot grant awarded to S.B. from the North Carolina Translational and Clinical Sciences Institute under Grant No. UL1TR002489. The authors would also like to thank Ms. Abby Wortman (Clinical and Lab Instructor for the Respiratory Care Program at South Dakota State University) for checking the scientific nomenclature on respiratory physiology used in this study.

## Author contributions

M.Y. = spatial discretization; computational simulations; post-processing; preliminary writing

M.M.H.A. = computational simulations; post-processing; literature review

A.M. = computational simulations

A.A.T.B. = computational simulations

A.T. = literature review

J.W. = flow experiments

W.D.B. = flow experiments

W.M.B. = UV-Vis spectroscopy

J.S.K. = CT segmentation with anatomical accuracy; digital reconstruction

A.C. = literature review; statistical inputs

J.R.P. = virological inputs; literature review; writing

S.B. = conceptualization and funding acquisition; study design; CT segmentation; digital reconstruction; data analysis; writing

## Additional information

The authors declare no competing interests.

## Data availability

All essential information is included in the article. Supplemental data (comprising anatomical geometries, computational simulation files, post-processing schemes, data spreadsheets) have been deposited to figshare, with DOI: 10.6084/m9.figshare.29363435.

